# Mapping the Clinical Trial Landscape in Anorexia Nervosa: A Registry-Based Analysis of Research Activity and Translational Gaps

**DOI:** 10.64898/2026.03.19.26348323

**Authors:** Bogdan Galusca, Natacha Germain, Manish Sarkar, Bérénice Gandit, Dimitrije Milunov, Kingsley Urakpo, Moutayam Khaddour, Soham Saha

**Affiliations:** TAPE Laboratory, University Jean Monnet, Saint Etienne; Endocrinology Unit, CHU Saint Etienne, Saint Etienne; MedInsights SAS, 24 Boulevard Saint-Jacques, 75014 Paris; NADDBio LLC, 245 First Street, Suite 1800, Cambridge, MA 02142-1292, United States

**Keywords:** Anorexia nervosa, clinical trials, explainable AI, large language models, retrieval augmented generators, feeding disorder, psychiatry, metabolism, eating behavior

## Abstract

**Background:** Anorexia nervosa (AN) is a severe psychiatric disorder associated with profound malnutrition, multisystem medical complications, and one of the highest mortality rates among mental illnesses. Despite decades of research into its biological and neurocognitive mechanisms, effective pharmacological treatments remain limited. While systematic reviews synthesize results from published studies, clinical trial registries offer a complementary perspective by capturing ongoing research efforts, discontinued studies, and emerging therapeutic strategies that may not yet be reflected in the published literature.

**Objective:** This study aimed to characterize the landscape of clinical research in AN by systematically analyzing studies registered on ClinicalTrials.gov.

**Methods:** We conducted a structured analysis of studies registered on ClinicalTrials.gov related to AN. Trial characteristics, including study design, intervention type, phase classification, geographic distribution, and recruitment status, were extracted and analyzed using an automated text-based classification pipeline.

**Results:** Nearly 400 studies investigating AN were identified over the past 25 years. Approximately 71% were classified as interventional studies; however, a large proportion were not associated with conventional clinical trial phases, suggesting that many registered trials correspond to mechanistic or exploratory investigations rather than therapeutic development programs. The geographic distribution of studies revealed a strong predominance of North America and Western Europe. A substantial proportion of trials were terminated or discontinued, highlighting the significant challenges associated with conducting interventional studies in this population. Observational studies generally included larger sample sizes than interventional trials.

**Conclusions:** Registry-based analyses provide valuable insights into the evolving landscape of clinical research in AN. Despite considerable scientific activity, important gaps remain between mechanistic knowledge and the development of therapeutic interventions. Understanding these gaps may help inform future translational research strategies aimed at improving treatment options for this severe disorder.

## Introduction

Anorexia nervosa (AN) is a severe psychiatric disorder characterized by persistent restriction of energy intake, intense fear of gaining weight, and disturbances in body image, leading to significantly low body weight relative to age, sex, developmental trajectory, and physical health (American Psychiatric Association, 2022). Beyond its psychiatric dimension, AN is also a condition of profound and often prolonged undernutrition that affects multiple physiological systems. Chronic energy deficiency leads to widespread medical complications including osteoporosis, endocrine alterations, cardiovascular abnormalities, and immune dysfunction (Estour et al., 2010; Galusca et al., 2006; Mehler and Brown, 2015; Misra and Klibanski, 2014a). These complications contribute to the particularly high mortality associated with AN, which remains one of the deadliest psychiatric disorders, with standardized mortality ratios substantially higher than those observed in most other mental illnesses (Arcelus et al., 2011).

The management of AN therefore requires a complex multidisciplinary approach combining psychiatric care, nutritional rehabilitation, and medical monitoring of complications related to severe malnutrition. Current treatment strategies rely largely on psychological and behavioral interventions aimed at restoring nutritional intake and addressing dysfunctional cognitive processes related to weight and body image (Treasure et al., 2020; Zipfel et al., 2015). However, treatment outcomes remain heterogeneous, and longitudinal studies indicate that approximately 20–25% of patients develop a chronic course characterized by persistent restrictive behaviors, repeated episodes of severe malnutrition, and long-term medical complications (Steinhausen, 2009).

One of the major challenges in the field is the absence of effective pharmacological treatments targeting the core mechanisms of AN. Unlike many other psychiatric or metabolic disorders, no medication has yet demonstrated consistent efficacy in improving the central symptoms of the disorder (Attia and Walsh, 2009; Treasure et al., 2020). Pharmacological approaches could theoretically target several aspects of AN pathophysiology, including dysregulation of appetite and satiety signaling, alterations in reward processing and decision-making related to food intake, and neuroendocrine adaptations to chronic starvation (Kaye et al., 2013; Turan et al., 2013). In addition, specific therapeutic strategies are needed to address somatic complications associated with severe and prolonged undernutrition, such as bone loss and endocrine dysfunction (Galusca et al., 2006; Mehler and Brown, 2015).

Over the past decades, considerable efforts have been devoted to investigating the biological mechanisms involved in the onset, maintenance, and progression of AN. In particular, increasing attention has been given to circulating peptides and hormones involved in appetite regulation, energy homeostasis, neuroendocrine signaling, and metabolic adaptation to starvation. These molecules have been explored both as potential contributors to disease mechanisms and as candidate biomarkers of disease severity, treatment response, or prognosis (Kaye et al., 2013; Monteleone and Maj, 2013; Prince et al., 2009). In a recent scoping review conducted by our group, we identified an extensive body of literature investigating circulating peptides in AN, encompassing more than two hundred molecules across several hundred studies (Doua et al., 2025). Despite this richness of mechanistic research, only a limited proportion of these candidate biological targets appear to have progressed toward clinical translation, either as therapeutic interventions or as clinically useful biomarkers.

Unlike systematic reviews that synthesize evidence from published studies, analyses based on clinical trial registries provide a complementary perspective on the research landscape. By examining registered protocols rather than published outcomes, such approaches capture ongoing investigations, discontinued or unsuccessful trials, and emerging therapeutic strategies that may not yet be visible in the published literature. Registry-based analyses also allow exploration of the global distribution of research activity and the diversity of methodological approaches currently implemented in the field.

In this context, the present study aimed to analyze studies registered on ClinicalTrials.gov focusing on AN. Using an automated text-based classification pipeline designed to extract and structure information from trial records, we sought to characterize the spectrum of interventions investigated, the biological targets explored, and the extent to which previously identified candidate mechanisms have translated into clinical research programs.

## Materials and methods

### Study design and overview

We developed an automated classification pipeline, called ClinForecast^TM^ system (trademarked with Paris RCS, INPI N° National: 26 5225815), to extract, structure and classify clinical trials data for systemic diseases. The system processes unstructured and semi-structured clinical trial records from ClinicalTrials.gov (Zarin et al., 2011) and applies a multi-agent generative and explainable artificial intelligence (xAI) architecture to classify key clinical entities according to predefined taxonomies. The methodology employs large language models (LLMs) with structured output enforcement, chain-of-thought reasoning, and domain-specific prompt engineering to achieve reproducible, classifications suitable for downstream pharmaceutical research. The ClinForecast^TM^ system was designed with a modular architecture to accommodate expansion across multiple organ systems and therapeutic areas. This methodology presently covers the expanded systems configuration, which encompasses hepatic, renal, endocrine, metabolic, hematologic, neurologic, immunologic and musculoskeletal disease categories (around 50000 trials information).

### Search criteria

The study selection process for identifying Anorexia nervosa (AN) trial studies was conducted through a systematic pipeline consisting of identification, screening, and inclusion phases (Page et al., 2021). Initially, 508 records were identified from the ClinicalTrials.gov register. The initial search criteria were the selection of trials with the following terms in the title, description and conditions mentioned in ClinicalTrials.gov: “Anorexia Nervosa” OR “Anorexia” OR “Anorectic” OR “Eating Disorder” OR “AN”. Before the formal screening process began, 27 duplicate records were removed, while no records were marked as ineligible by automation tools or removed for other reasons at this stage. This resulted in 481 trial records moving forward to the screening phase (**Figure 2**).

During the screening stage, all 481 records were processed using an API extraction protocol. No trials were excluded during this initial automated screening, as all 481 records met the requirement of containing “anorexia nervosa” or “AN” in the title and description. Consequently, 481 reports were sought for retrieval, and all 481 were successfully retrieved for a full eligibility assessment. The final eligibility assessment utilized RAG/LLM fine-tuned models and ClinForecast tools for filtering. During this stage, 81 reports were excluded: 35 based on inclusion criteria addressing the “Anorexia nervosa” and/or “AN” condition, and 46 based on exclusion criteria where trials did not include the “Anorexia nervosa” and/or “AN” condition. Following these exclusions, a total of 400 studies were included in the final review. The selection follows the PRISMA guidelines (Page et al., 2021) and final results are presented in **Supplementary table 1**.

### Disease profile

#### Disease-specific psychopathology and diagnostic subtypes

Endpoints in this domain were established to capture distinct diagnostic entities, core psychopathological features, and subtype-specific mechanisms within anorexia nervosa (AN) requiring tailored therapeutic strategies based on illness subtype, severity grading, and phenotypic presentation. Diagnostic categorization utilized the Diagnostic and Statistical Manual of Mental Disorders, Fifth Edition (DSM-5) criteria to stratify patients into restricting or binge-eating/purging subtypes (Call et al., 2013). Severity grading was defined by body mass index (BMI) thresholds, ranging from mild (BMI ≥ 17 kg/m²) to extreme (BMI < 15 kg/m²). Atypical AN was identified by significant weight loss and psychological morbidity in the absence of an absolute low BMI (Sawyer et al., 2016). Core psychopathology was quantified by assessing the fear of weight gain, body image distortion, ego-syntonicity, and the neurobiologically mediated severity of anosognosia. Illness trajectories were classified by duration and age of onset, while trauma comorbidity profiling was utilized to document the presence or absence of co-occurring post-traumatic stress disorder, serving as a cross-reactive immunologic material equivalent.

#### Survival and disease progression metrics

Survival and disease progression were evaluated using hierarchical measures of mortality risk, chronicity, relapse, and the achievement of recovery milestones. Key metrics included overall survival, standardized mortality ratios, and the time required for weight restoration to at least 90% of ideal body weight or an age-adjusted BMI percentile. The study monitored relapse-free survival and time to relapse, defined by weight loss, the return of restrictive behaviors, or re-hospitalization. Physiological recovery was tracked via the time to resumption of menses and the normalization of vital signs, including heart rate, blood pressure, and temperature. Healthcare utilization and disease chronicity were assessed through the duration and frequency of hospitalizations, 6- and 12-month post-discharge readmission rates, and the transition rate from acute to severe-enduring AN. Furthermore, the protocol tracked the time to loss of functional independence, the rate of progression to chronic multimorbidity lock-in, and the time to severe cardiac events such as arrhythmias or cardiac arrest.

#### Biochemical and metabolic control

Laboratory-based assessments of metabolic homeostasis, nutritional status biomarkers, and biochemical changes were utilized to reflect disease severity and treatment response. Routine monitoring included serum electrolytes, blood glucose levels, hypoglycemic episode frequency, and complete blood counts to detect bone marrow suppression sequelae such as leukopenia, anemia, and thrombocytopenia. Hepatic transaminases were measured to identify starvation-induced hepatopathy, while albumin and prealbumin served as nutritional status markers. The evaluation encompassed lipid profiles to detect paradoxical hypercholesterolemia, renal function under volume depletion via blood urea nitrogen and creatinine, lactate levels, and acid-base balance alterations resulting from purging or laxative abuse. Amylase levels were tracked as a biomarker for parotid hyperamylasemia in the purging subtype. Disease-specific biomarkers analyzed included appetite and satiety hormones (leptin, ghrelin, and peptide YY), insulin-like growth factor 1 as a marker of growth hormone resistance, and cortisol levels reflecting hypothalamic-pituitary-adrenal axis hyperactivity. Thyroid panels, gonadal hormones, gonadotropins, and bone turnover markers (osteocalcin, CTX, P1NP) were also quantified. Thiamine levels were assessed for Wernicke-Korsakoff risk, and microbiome diversity indices were explored as emerging biochemical markers.

#### Genetic, epigenetic, and neurobiological biomarkers

Molecular and neuroimaging markers were employed to guide patient characterization, predict treatment response, and define disease vulnerability profiles. Genetic assessments incorporated polygenic risk scores for AN, genome-wide association study risk loci, and heritability estimates. Genotype-phenotype correlations were evaluated to understand the overlap between metabolic and psychiatric genetics, supplemented by detailed family histories of eating disorders, leanness, obsessive-compulsive disorder, and anxiety. Epigenetic modifications, specifically starvation-induced DNA methylation patterns and markers of transgenerational epigenetic transmission, were analyzed alongside gene-environment interaction profiling and autoimmune-related genetic loci overlap. Neurobiological characterization utilized structural magnetic resonance imaging to detect grey and white matter volume changes. Functional MRI was employed to assess activation patterns in fronto-striatal reward circuits, the extra-striate body area, and the fusiform body area, while saccadic eye movement abnormalities were monitored as a behavioral proxy for neural dysfunction.

#### Nutritional rehabilitation and refeeding outcomes

The efficacy and safety of nutritional restoration were evaluated through precise monitoring of weight trajectories, dietary adequacy, and physiological tolerance during inpatient and outpatient treatment. Key metrics included the rate of weekly weight gain, achievement of target BMI percentiles, and the successful escalation of caloric prescriptions. Refeeding safety was rigorously assessed by monitoring the incidence and severity of refeeding syndrome, specifically tracking hypophosphatemia, hypomagnesemia, hypokalemia, and phosphate nadirs (Friedli et al., 2018). Clinical interventions required to achieve nutritional goals, such as the necessity for nasogastric or naso-jejunal feeding, medical food tolerance, and the time required to transition to independent oral feeding, were documented. Behavioral nutritional outcomes included oral caloric intake adequacy, dietary variety, meal completion rates, and tolerance to caloric density escalation. Finally, metabolic recovery was measured through body composition analysis, distinguishing between fat and lean mass restoration, and the normalization of resting energy expenditure.

#### Psychotherapeutic and behavioral intervention outcomes

Treatment response metrics were systematically collected for evidence-based psychological and behavioral therapies to gauge engagement, adherence, and the achievement of therapeutic milestones. Outcomes were evaluated across multiple modalities, including Family-Based Treatment, Enhanced Cognitive Behavioral Therapy, the Maudsley AN Treatment for Adults, Specialist Supportive Clinical Management, and Focal Psychodynamic Therapy. Adjunctive cognitive interventions, such as Cognitive Remediation Therapy and Cognitive Bias Modification, were assessed for their impact on improving cognitive flexibility. General treatment metrics included therapy completion versus premature dropout rates, motivational enhancement scores, and the quality of the therapeutic alliance. The study also tracked the progression through stepped-care models, the rates and outcomes of involuntary treatment, and the time required in treatment before observing clinically significant improvements.

#### Neuroendocrine and hormonal restoration

The protocol assessed the recovery of endocrine axes, hormonal normalization, and the resolution of starvation-induced endocrine adaptations. Recovery of the hypothalamic-pituitary-gonadal axis was monitored via the restoration of luteinizing hormone pulsatility, normalization of estradiol and testosterone levels, and the ultimate resumption of menstrual cyclicity, noting the specific age-adjusted BMI percentile threshold at which menses returned (Misra and Klibanski, 2014b). Hypothalamic-pituitary-adrenal axis normalization was determined by the restoration of diurnal cortisol rhythms, while thyroid axis recovery was marked by the resolution of euthyroid sick syndrome. Normalization of insulin-like growth factor 1 indicated the resolution of growth hormone resistance, which was correlated with growth velocity recovery and Tanner stage pubertal progression in pediatric and adolescent cohorts. Furthermore, the recovery trajectories of metabolic hormones, including leptin, adiponectin, resistin, ghrelin, and peptide YY, were continuously tracked.

#### Neurological, neurodevelopmental and cognitive outcomes

Central and peripheral nervous system function, cognitive recovery, and neurodevelopmental impacts were comprehensively assessed both before and following weight restoration. Brain structural restoration, specifically the reversal of pseudoatrophy, was quantified using MRI volumetrics for grey and white matter. Neuropsychological batteries evaluated cognitive flexibility using the Wisconsin Card Sorting Test and Trail Making Test B, while central coherence was assessed via the Rey-Osterrieth Complex Figure Test (Tchanturia et al., 2012). Additional cognitive domains evaluated included set-shifting capacity, executive function, attention, concentration, and processing speed. The resolution of anosognosia and visual body processing normalizations were tracked using functional MRI paradigms targeting the extra-striate and fusiform body areas. Autonomic nervous system recovery was measured via heart rate variability, and saccadic eye movements were monitored for normalization. For pediatric patients, developmental milestones were assessed using appropriate scales, alongside evaluations of post-treatment academic functioning and markers of neural plasticity during recovery.

#### Organ function and multi-system complications

The assessment of end-organ damage, disease-related medical complications, and multi-system involvement was systematically integrated into the evaluation. Cardiovascular monitoring included heart rate, blood pressure, QTc intervals, echocardiographic structural assessments, and the screening for postural tachycardia. Skeletal health was quantified using dual-energy X-ray absorptiometry to assess bone mineral density, grade osteopenia or osteoporosis, and track fracture incidence (Fazeli and Klibanski, 2014). Gastrointestinal evaluations screened for gastroparesis via gastric emptying studies, superior mesenteric artery syndrome, hepatic steatosis, and constipation severity. Renal function and hematological integrity were evaluated through glomerular filtration rates, complete blood counts to detect bone marrow suppression, and electrolyte panels. Dermatological and dental manifestations, such as lanugo, acrocyanosis, Russell’s sign, and perimylolysis, were documented. The protocol also monitored thermoregulatory deficits, pulmonary risks including pneumomediastinum in purging subtypes, and reproductive structural recovery measured by ovarian volume and uterine size via ultrasound.

#### Quality of life and patient-reported outcomes

Patient-centered measures were employed to evaluate the subjective impact of the disease and treatment on daily functioning, well-being, and global recovery. Standardized instruments, including the Eating Disorder Quality of Life questionnaire, EQ-5D-5L utility scores, the SF-36 health survey, and the Eating Disorder Examination Questionnaire, were utilized to capture clinical impairment and symptom burden. Psychological comorbidities and physical well-being were further quantified using the Patient Health Questionnaire-9 for depression, the Generalized Anxiety Disorder-7 scale, and PROMIS measures for physical function, fatigue, and pain. Beyond individual symptom scales, the study assessed broader psychosocial impacts through caregiver burden assessments, work or school attendance, social functioning, and independence in activities of daily living. Subjective clinical remission and routine healthcare utilization patterns, including emergency department visits, were also recorded as holistic indicators of recovery.

#### Symptom burden and clinical manifestations

Eating disorder-specific symptoms, compensatory behaviors, and acute clinical presentations were meticulously monitored throughout the study period. Core symptom burden was quantified using the Eating Disorder Examination interview scores and Eating Disorder Inventory-3 subscales. The severity of caloric restriction, rigid food rules, binge-eating episodes, and purging behaviors - including self-induced vomiting and the misuse of laxatives or diuretics - were continuously tracked. Additional behavioral manifestations assessed included the frequency and compulsivity of exercise using the Compulsive Exercise Test, body checking and avoidance behaviors, and body image disturbance measured by the Body Shape Questionnaire. The evaluation also captured ritualistic eating behaviors, deficits in interoceptive awareness, fear of food, meal-related distress, night eating syndromes, and screening for insulin misuse or diabulimia in comorbid type 1 diabetes populations.

#### Psychiatric comorbidity and emotion regulation

The interplay between eating pathology and co-occurring psychiatric conditions, emotion dysregulation, and trauma was rigorously evaluated. The prevalence and severity of comorbid diagnoses, including depression, anxiety, obsessive-compulsive disorder, post-traumatic stress disorder, and substance use, were assessed using validated psychometric scales such as the Beck Depression Inventory-II, the State-Trait Anxiety Inventory, and the Yale-Brown Obsessive-Compulsive Scale. Trauma history and symptom severity were evaluated via the Childhood Trauma Questionnaire and the Clinician-Administered PTSD Scale for DSM-5. Furthermore, the assessment of psychological endophenotypes and emotional functioning incorporated the Difficulties in Emotion Regulation Scale, the Toronto Alexithymia Scale, and measures of perfectionism and harm avoidance. Routine screenings for self-harm and suicidality were conducted utilizing the Columbia Suicide Severity Rating Scale, alongside evaluations for autism spectrum traits and specific personality temperament profiles.

#### Pharmacological intervention and safety monitoring

The efficacy, safety, and tolerability of pharmacological interventions utilized in the management of AN were continuously monitored. Medication adherence and target symptom responses were evaluated for atypical antipsychotics, specifically assessing olanzapine for weight gain and anxiety reduction, as well as for antidepressants and anxiolytics in weight-restored patients. The protocol also tracked outcomes related to hormonal therapies, cannabinoid receptor agonists, zinc supplementation, and novel experimental pharmacotherapies such as psilocybin or oxytocin. Comprehensive safety monitoring was mandated, encompassing adverse event incidence, QTc interval prolongation on psychotropic medications, metabolic shifts induced by antipsychotics, and potential drug-drug interactions. Strict clinical guidelines were observed, including the contraindication of thyroid hormone replacement in the context of euthyroid sick syndrome and mandatory thiamine repletion prior to glucose administration to prevent neurological sequelae.

#### Microbiome, immune and novel biological targets

Endpoints measuring emerging biological targets focused on gut-brain axis function, immune markers, and novel pathophysiological mechanisms. Gut microbiome diversity was quantified using Shannon and Chao1 indices, alongside structural shifts in the Firmicutes-to-Bacteroidetes ratio and variations in short-chain fatty acid profiles. Gut permeability was assessed via zonulin and lipopolysaccharide levels, while systemic and central inflammation were monitored through inflammatory cytokine profiles (including IL-6, TNF-α, and CRP) and neuroinflammation markers. The evaluation extended to the tryptophan-kynurenine pathway, autoimmune anti-neuronal antibodies, and gut-brain axis signaling markers such as vagal tone and enteric serotonin. Central nervous system targets included neurotransmitter metabolite profiles, epigenetic clock measurements, and circadian rhythm markers. Additionally, functional neuroimaging captured reward circuit responses during food cue paradigms, fronto-striatal connectivity, and amygdala reactivity to trauma-related stimuli.

#### Other/unclassified

Endpoints that did not distinctly align with the primary clinical and biological categories described above were classified separately to ensure comprehensive data capture. This included isolated measures of specific drug efficacy, pharmacokinetic and pharmacodynamic profiles, and exclusive reports of safety, tolerability, or adverse events. Furthermore, isolated metrics pertaining solely to protocol compliance, adherence measures, health economic evaluations, cost-effectiveness endpoints, and implementation or service delivery outcomes were aggregated within this domain for supplementary analysis.

### Data Sources

#### Primary clinical trial data

Clinical trial records were obtained from ClinicalTrials.gov, a publicly accessible registry maintained by the U.S. National Library of Medicine. Data was retrieved via the ClinicalTrials.gov Application Programming Interface (API) version 2, which returns structured JSON-formatted records. Key fields extracted for classification included: (1) study title, brief summary, and detailed description; (2) eligibility criteria; (3) intervention/treatment arms; (4) primary and secondary outcome measures; and (5) sponsor and collaborator information.

### Classification framework

The ClinForecast^TM^ system extracts and classifies the following entity categories from each clinical trial record:

**Table 1.** Clinical Trial Entities Subject to Automated Classification.

| Entity Category | Description and Classification Scope |
| --- | --- |
| Interventions | Drug modalities and technology platforms classified by mechanism of action |
| Endpoints | Primary and secondary outcomes classified by pathophysiology domain and efficacy/biomarker type |
| Eligibility Criteria | Inclusion and exclusion criteria with patient population context |
| Trial Phase/Status | Development stage (Phase I-IV) and recruitment status |
| Sponsors | Lead sponsor categorization by organizational type (Industry, Academia, Other) |

#### Drug and technology modality classification

Therapeutic interventions were classified using a hierarchical decision-tree taxonomy derived from contemporary pharmaceutical development conventions (Yamaguchi et al., 2021). The classification schema defines six primary molecule types:

1. Small molecules (<1 kDa molecular weight, chemically synthesized)
2. Protein-based biologics (monoclonal antibodies, fusion proteins, enzymes)
3. Nucleic acid therapeutics (antisense oligonucleotides, siRNA, mRNA, CRISPR)
4. Cell-based therapies (CAR-T, stem cell therapies)
5. Gene therapies (AAV vectors, lentiviral vectors)
6. Hybrid modalities (antibody-drug conjugates, radioligand therapies)

Within each primary category, secondary technology modality classifications (>20 specific technologies) provide granular characterization, including bispecific antibodies, proteolysis-targeting chimeras (PROTACs), autologous versus allogeneic cell therapies, and various delivery platforms. The decision-tree logic was encoded as structured prompt instructions requiring sequential feature evaluation before final classification assignment.

#### Endpoint pathophysiology classification

Clinical trial endpoints were classified according to organ-system-specific pathophysiology domains. The classification framework distinguished between direct clinical efficacy measures (how patients feel, function, or survive) and biomarker endpoints (indicators of biological response), consistent with FDA-NIH Biomarkers, Endpoints, and other Tools (BEST) resource definitions (National Institutes of Health (US), 2025). Endpoint categories were contextualized to each therapeutic area. For example, within the metabolic systems configuration, classification categories include:

- Disease-specific biology (e.g., tumor-specific biology for oncology, metabolic pathway markers for inborn errors of metabolism)
- Survival and disease progression endpoints (overall survival, progression-free survival)
- Radiographic and imaging response criteria (RECIST, imaging biomarkers)
- Molecular and genetic biomarkers
- Patient-reported outcomes and quality of life measures
- Safety and tolerability endpoints

The modular system architecture allows organ-system-specific endpoint taxonomies to be defined independently, with configuration files specifying the applicable classification categories for each therapeutic area.

### Classification Methodology

#### Multi-Agent LLM Architecture

The classification pipeline employs a multi-agent architecture integrating OpenAI GPT-series models (GPT-5, GPT-5.2). Specialized classification agents were instantiated for each entity type, with distinct agent configurations defined in the system architecture:

- Drug mechanism of action agent: Classifies interventions by modality and technology
- Endpoint pathophysiology agent: Classifies outcomes by disease domain
- Eligibility criteria agent: Extracts and structures inclusion/exclusion criteria

Each agent receives domain-specific instructions from prompt templates that define the agent persona (role description), classification rules, decision logic, and output schema requirements.

#### Chain-of-Thought reasoning and structured output enforcement

All classification prompts mandate chain-of-thought reasoning before output generation (Wei et al., 2022). Each classification request requires the model to produce:

1. A reasoning field articulating the logical basis for classification
2. A confidence score (categorical: high, medium, low or continuous: 0.0-1.0)
3. The final classification label(s) conforming to the predefined taxonomy

Output schema compliance is enforced through Pydantic data models (Pulla et al., 2024) that define rigid type constraints for all classification outputs. The LLM is constrained to generate valid JSON conforming to these schemas, ensuring structural compliance with expected data types and enumerated value sets. This approach eliminates parsing errors and guarantees downstream data integrity.

#### Decision tree logic encoding

Complex classification logic, particularly for drug modality assignment, was encoded as textual decision trees within prompt instructions. The decision tree structure forces the LLM to evaluate classification criteria sequentially. For drug modality classification, the decision path includes questions such as: (1) Is the molecular weight less than 1 kDa? (2) Is the entity protein-based? (3) Does the mechanism involve nucleic acid delivery? This approach aligns LLM reasoning with expert pharmaceutical classification practice.

#### Intelligent caching for consistency and efficiency

To ensure classification consistency across the dataset and reduce computational costs, the system implements intelligent caching at two levels:

##### Entity-level caching

Drugs and sponsors are cached by normalized name. Once an entity is classified, subsequent occurrences retrieve the cached classification, ensuring consistency across all trials involving that entity.

##### Content-based caching

Whole-trial inputs are hashed to detect duplicate or near-duplicate records, preventing redundant classification and enabling efficient reprocessing when only portions of the dataset require reclassification.

### Validation and Quality Control

#### Schema compliance validation

All classification outputs were validated against predefined Pydantic schemas that enforce type constraints, permitted enumeration values, and required field presence. Schema validation achieves 100% structural compliance, as outputs failing validation are rejected and flagged for review. Unit testing verifies correct loading of organ-system-specific prompts and configuration files across all system modules.

#### Confidence-based quality flagging

Classification outputs include model-generated confidence scores. Low-confidence classifications (confidence < 0.6 or categorical *Low*) are automatically flagged for human review. This approach balances automation efficiency with quality assurance for ambiguous or borderline cases, consistent with recommendations for human-in-the-loop AI systems in clinical research (Terranova et al., 2023).

#### Manual quality control review

A subset of classifications underwent manual review by domain experts to validate accuracy. Eligibility criteria classifications, which involve particularly nuanced clinical interpretation, were subject to structured human review with documented acceptance/rejection decisions. Classification errors identified during manual review were used to iteratively refine prompt instructions and decision logic.

#### Deduplication and data integrity

Prior to analysis, the dataset was processed through deduplication procedures to identify and remove duplicate trial records. Deduplication logic accounts for trial version updates, registry cross-listings, and re-registrations that may create apparent duplicates in the source data.

### Limitations

Several methodological limitations warrant consideration. First, the classification pipeline relies on LLM inference, which may introduce stochastic variability in edge cases despite prompt engineering and schema enforcement. Second, classifications are dependent on the completeness and accuracy of source data in ClinicalTrials.gov; trials with incomplete or ambiguous descriptions may yield uncertain classifications. Third, the taxonomies employed represent expert-derived consensus categories but may not capture all nuances of rapidly evolving therapeutic modalities. Fourth, validation against manually curated gold-standard datasets is ongoing, and accuracy metrics should be interpreted as preliminary until larger validation studies are completed. Finally, as with all AI-assisted classification systems, periodic prompt and model updates may be required to maintain accuracy as new therapeutic modalities and clinical terminology emerge. A potential limitation of this analysis is that it relied primarily on ClinicalTrials.gov, which represents only one component of the global clinical trial registration landscape. Several studies have shown that a substantial proportion of clinical trials are registered in other national or regional registries integrated within the WHO International Clinical Trials Registry Platform (ICTRP), including the European Union Clinical Trials Register, the Chinese Clinical Trial Registry, and the Clinical Trials Registry of India. Consequently, analyses restricted to ClinicalTrials.gov may underestimate the contribution of studies conducted in regions where local registries are more commonly used, potentially introducing a geographic registration bias (Banno et al., 2019; DeVito et al., 2020; Viergever and Li, 2015; Zarin et al., 2016).

## Results

### Data extraction and enrichment pipeline sifts and classifies AN clinical trials

The initial data ingestion for the ClinForecast^TM^ pipeline included a repository of clinical trials targeting AN, totaling 508 trials. Following a relevance assessment against established AN benchmark, 27 trials were excluded, leaving a final cohort of 481 trials for initial analysis. We extracted the final list considering the inclusion/exclusion criteria and ended up with 400 trials for the following analysis (**Figure 1**). The trials subsequently underwent advanced classification using a RAG/LLM approach, followed by the development of hierarchical logic and quantitative enrichment confirmed by expert validation. Data was then processed through extraction and transformation-load preparation into a comprehensive data enrichment pipeline. This multidimensional pipeline facilitated distinct analytical pathways, evaluating trial phases, current status, and failure reasoning, including specific categorization for the COVID-19 era. Additional operational analyses captured expected timelines, duration metrics per phase, sponsor breakdowns between industry and academia and granular patient enrollment data comparing healthy volunteers to specific patient populations. On the clinical side, the pipeline differentiated observational from interventional trials, identifying true interventional studies involving distinct therapeutic targets while also classifying eligibility criteria, disease stage relevance, and organ-related co-morbidity profiles. Trial interventions were deeply phenotyped by analyzing drug mechanisms of action (MoA), technology types and mechanistic classes (**Supplementary table 1**). Concurrently, pathophysiology analyses categorized both primary and secondary endpoints into localized pathophysiological clusters. These clinical and mechanistic insights directly fed into a terminal classification of safety, efficacy and biomarker endpoints. To finalize the dataset, all distributed data streams were synthesized through sequential consolidation, post-processing and homogenization steps (**Supplementary table 1**). The reliance on advanced RAG/LLM classification and expert validation demonstrates the growing complexity of parsing clinical trial registries. It highlights that raw trial data requires significant enrichment and standardization before meaningful conclusions can be drawn about disease stage pathophysiology and expected timelines.

**Figure 1:**
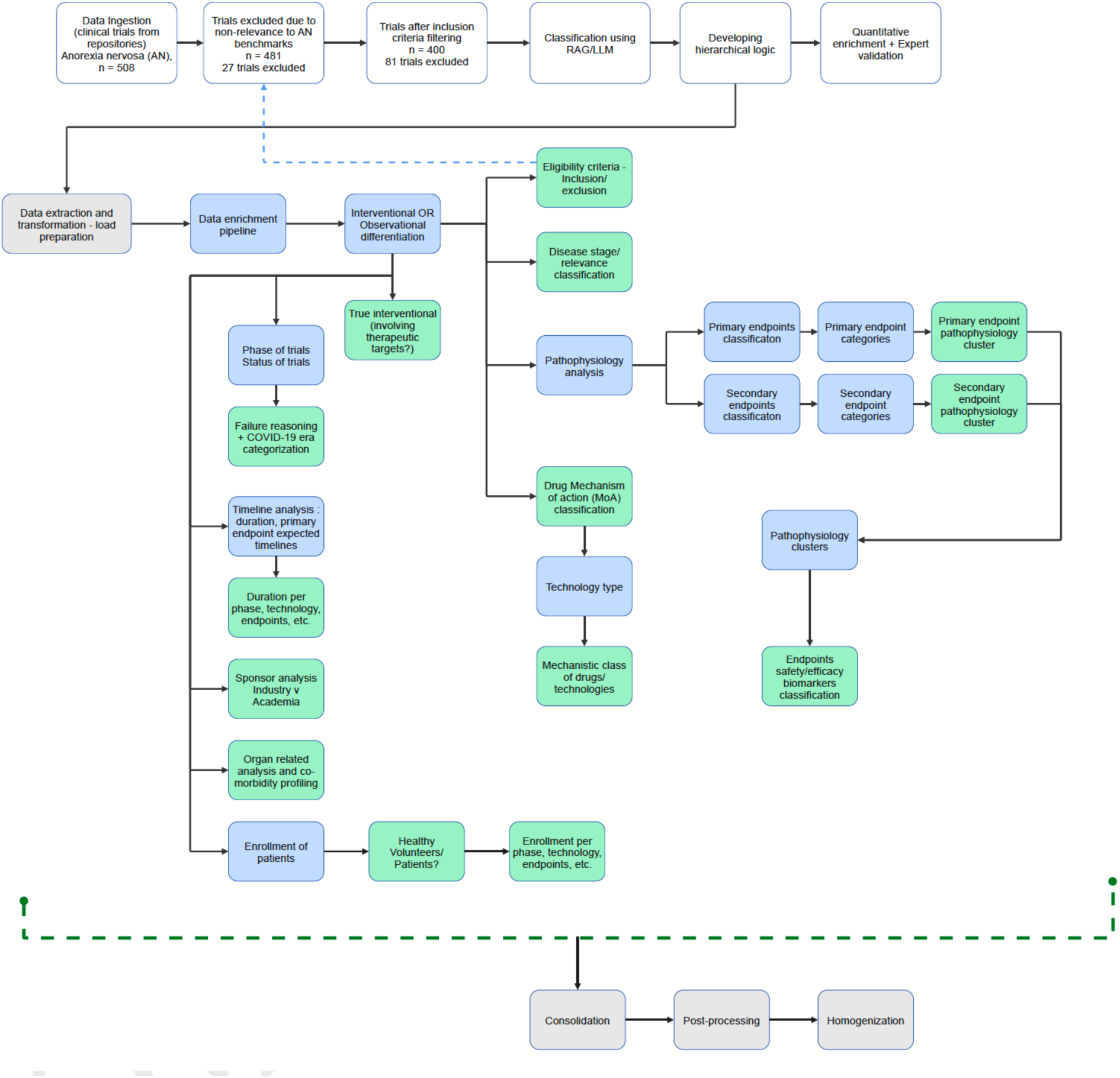
Overview of the ClinForecast^TM^ data processing and enrichment pipeline for AN clinical trial. The flowchart depicts the end-to-end methodology, beginning with the ingestion of 508 clinical trials from data repositories. After excluding trials due to non-relevance to AN benchmark or non-existence in inclusion criteria (leaving n=400), the core data enrichment pipeline systematically extracts and categorizes trial characteristics across multiple parallel domains following initial load preparation. Key analytical branches include operational metrics such as trial phase and status, timeline and duration analysis, sponsor categorization, and patient enrollment profiles. The clinical profiling pathways differentiate trial design (interventional vs. observational) and detail the classification of eligibility criteria, disease stage and pathophysiology. Furthermore, primary and secondary endpoints are mapped into specialized pathophysiological clusters, alongside drug mechanism of action (MoA) classification.

**Figure 2.**
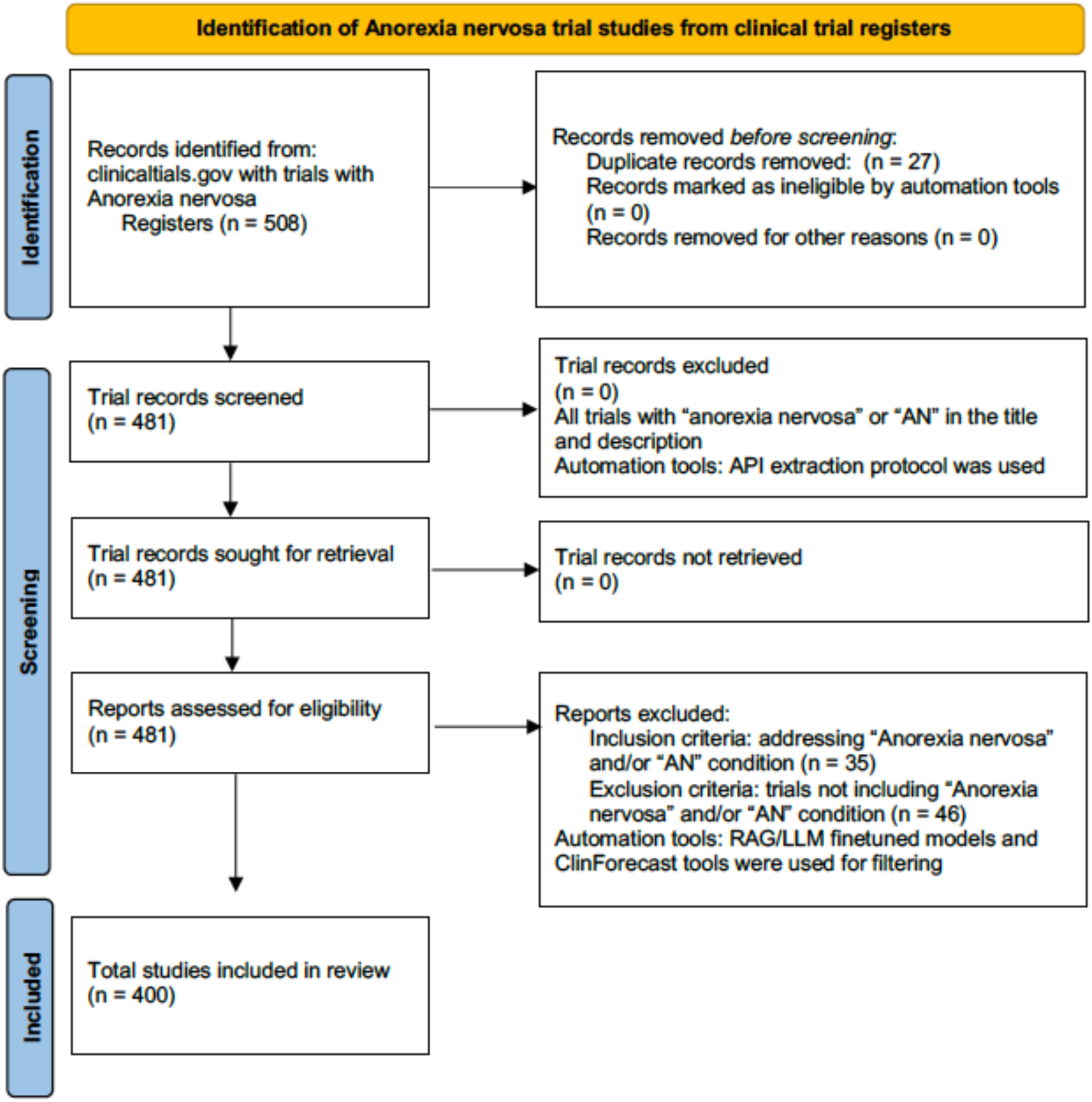
Identification of Anorexia nervosa trial studies from the clinical trial register including the criteria from included and excluded studies. (A) Distribution of study types, comparing observational to interventional designs across the total cohort of 400 trials. (B) Longitudinal trends demonstrating the number of study types per year. (C) Proportional breakdown of the clinical trial phases within the dataset. (D) Categorization of current study status, detailing active, completed, unknown, and failed trials. (E) Annual timeline showing the distribution of trial status

### Clinical trial landscape characterized by phase 2 dominance, recruitment challenges, and western geographic centralization

An analysis of the 400 identified clinical trials reveals a strong preference for interventional (71.5%) over observational study designs (**Figure 3A, Supplementary table 1**), with interventional trials steadily increasing in volume since the year 2000 (**Figure 3B**). The developmental pipeline is heavily concentrated in mid-stage research. The vast majority of CT phase was not available (75.96%), indicating incomplete or partial completeness in the execution of the trials (**Figure 3C**). Phase 2 trials accounted for 8.71% of the trials. Conversely, late-stage studies remain sparse, with Phase 3 and Phase 4 trials accounting for only 3.48% and 2.09% of the clinical landscape, respectively (**Figure 3C**). In terms of operational outcomes, nearly half of the cohort has successfully completed the study protocol (n=190) (**Figure 3D**), and an analysis of the temporal outcomes reveal that these studies were largely completed in the last 10 years (2015 to 2022), and new studies have started recruiting participants from 2022 onwards (**Figure 3E**). However, an analysis of failed trials, those categorized as withdrawn, terminated, or suspended, indicates that patient recruitment is the most significant hurdle, responsible for 45.16% of trial failures (**Figure 3F**). Secondary failure reasons were tied to principal investigator (PI) displacement (16.13%) and disruptions caused by COVID-19 (16.13%). Geographically, the distribution of trial sites is highly centralized in Western countries (**Figure 3G**). Europe hosts the highest number of overall trial sites at 247, closely followed by North America with 198 sites, while Asia (23 sites) and the Asia/Pacific region (1 site) are notably underrepresented (**Figure 3H**). At the individual country level, the United States leads global participation with 181 sites, followed by prominent European hubs including France (90 sites) and Italy (29 sites) (**Figure 3I**). Canada (23 sites), United Kingdom (23 sites), Sweden (21 sites), Denmark (17 sites), Spain (16 sites), Germany (15 sites) and China (11 sites) are other countries reported to be conducting AN trials (**Figure 3I**).

**Figure 3.**
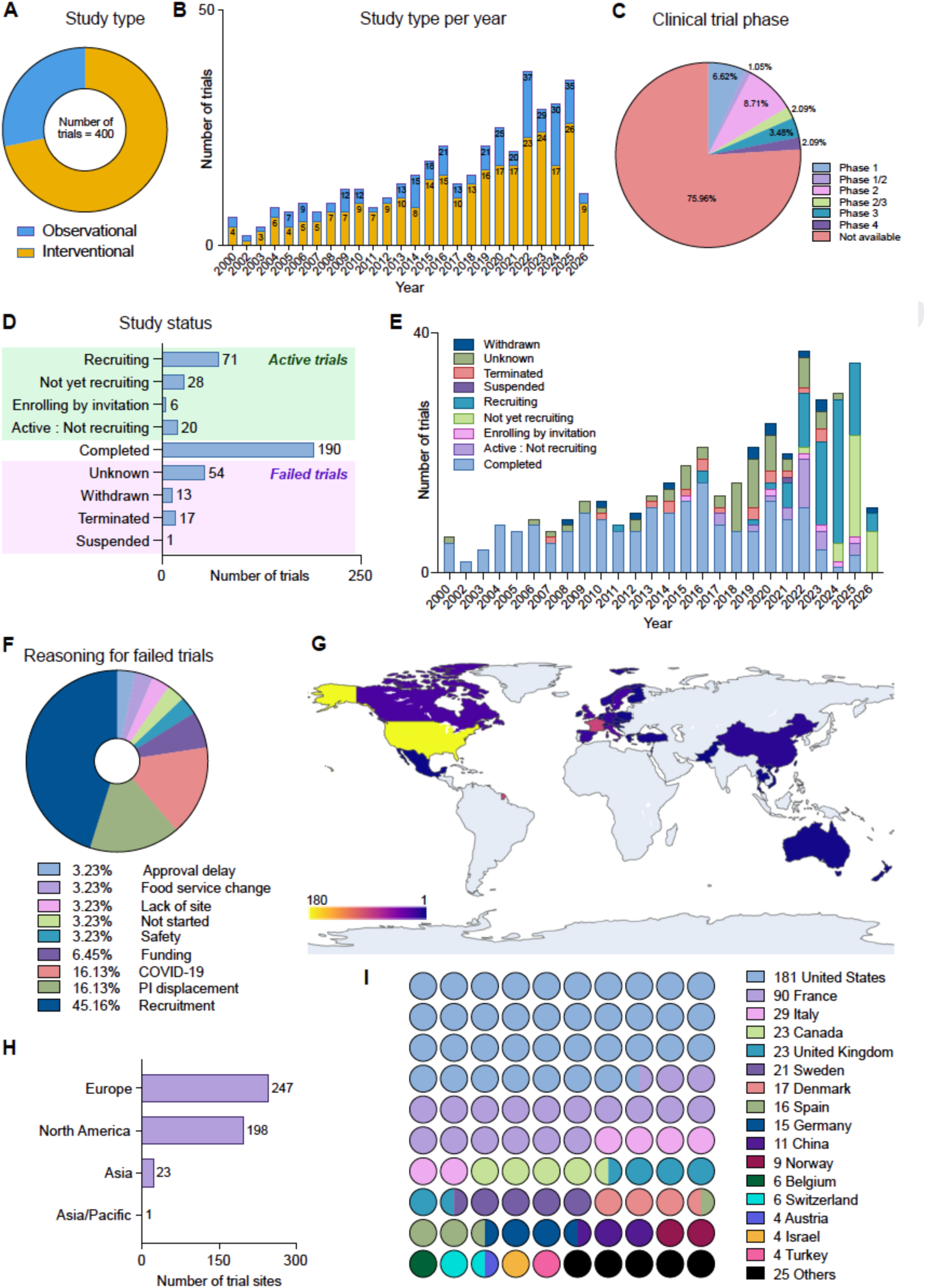
Landscape and characteristics of AN clinical trials. (A) Distribution of study types, comparing observational to interventional designs across the total cohort of 400 trials. (B) Longitudinal trends demonstrating the number of study types per year. (C) Proportional breakdown of the clinical trial phases within the dataset. (D) Categorization of current study status, detailing active, completed, unknown, and failed trials. (E) Annual timeline showing the distribution of trial status from 2000 to 2026. (F) Quantitative reasoning for failed trials (encompassing withdrawn, terminated, and suspended studies). (G) Global heatmap detailing the geographic distribution of trial sites. (H) Aggregate number of trial sites distributed by major geographic regions. (I) Specific breakdown of the number of trial sites hosted by the top participating countries globally.

### Efficacy and behavioral outcomes dominate the endpoint amidst fluctuating enrollment sizes

An in-depth analysis of clinical trial endpoints reveals a distinct prioritization of efficacy over safety or pharmacokinetic profiles in AN research (**Figure 4A, Supplementary table 1**). When mapping the hierarchical flow of trial objectives, “efficacy” overwhelmingly dominates the primary endpoint categories for both interventional and observational studies (**Figure 4B**). “Mixed” endpoint categories and “safety” categories were also observed, but largely for interventional studies. Further sub-stratification into endpoint classifications demonstrates that “clinical efficacy” drives the vast majority of trial volume (>300 trials), distantly followed by “imaging biomarker” and “molecular biomarker” metrics (**Figure 4C**). It is to be noted that “genetic/epigenetic biomarkers”, “pharmacokinetics” and “laboratory safety” remains low in this sub-stratification of endpoints (**Figure 4C**), indicating that the scope of AN molecular and metabolism-driven therapies remains large. At the most granular level of endpoint details, trial objectives are heavily concentrated on three core pathophysiological and behavioral domains: “eating behavior”, “neuropsychology”, and “metabolic regulation” (**Figure 4D**). While this study classifies a significant number of trial studies into “biomarker” driven endpoints, its classification into interventional frameworks warrants careful consideration. The clinical utility of such a marker is most pronounced when utilized as a tool for patient stratification - specifically in identifying cohorts most likely to benefit from therapeutic or behavioral interventions relevant to AN. Longitudinal tracking of average patient enrollment in interventional trials from 2000 to 2025 highlights significant temporal fluctuations depending on the endpoint targeted. While trials focusing on clinical efficacy and clinical safety maintain consistent baseline enrollment numbers, notable spikes are evident; for instance, trials classified with “multiple” or safety-focused endpoints saw a sharp increase in average patient enrollment size around 2019 (**Figure 4E**). The trials were also categorized into several endpoint details such as behavior, neuropsychology, neurology, eating behavior, metabolic regulation, molecular and skeletal-metabolic regulation, indicating the system that might be evaluated in the trials. When analyzing the average patient enrollment by specific endpoint details, trials measuring “neuropsychology” and “behavior” achieved notably high average enrollment peaks in 2011 and 2015, respectively, indicating periods of large-scale behavioral interventions within the field (**Figure 4F**).

**Figure 4.**
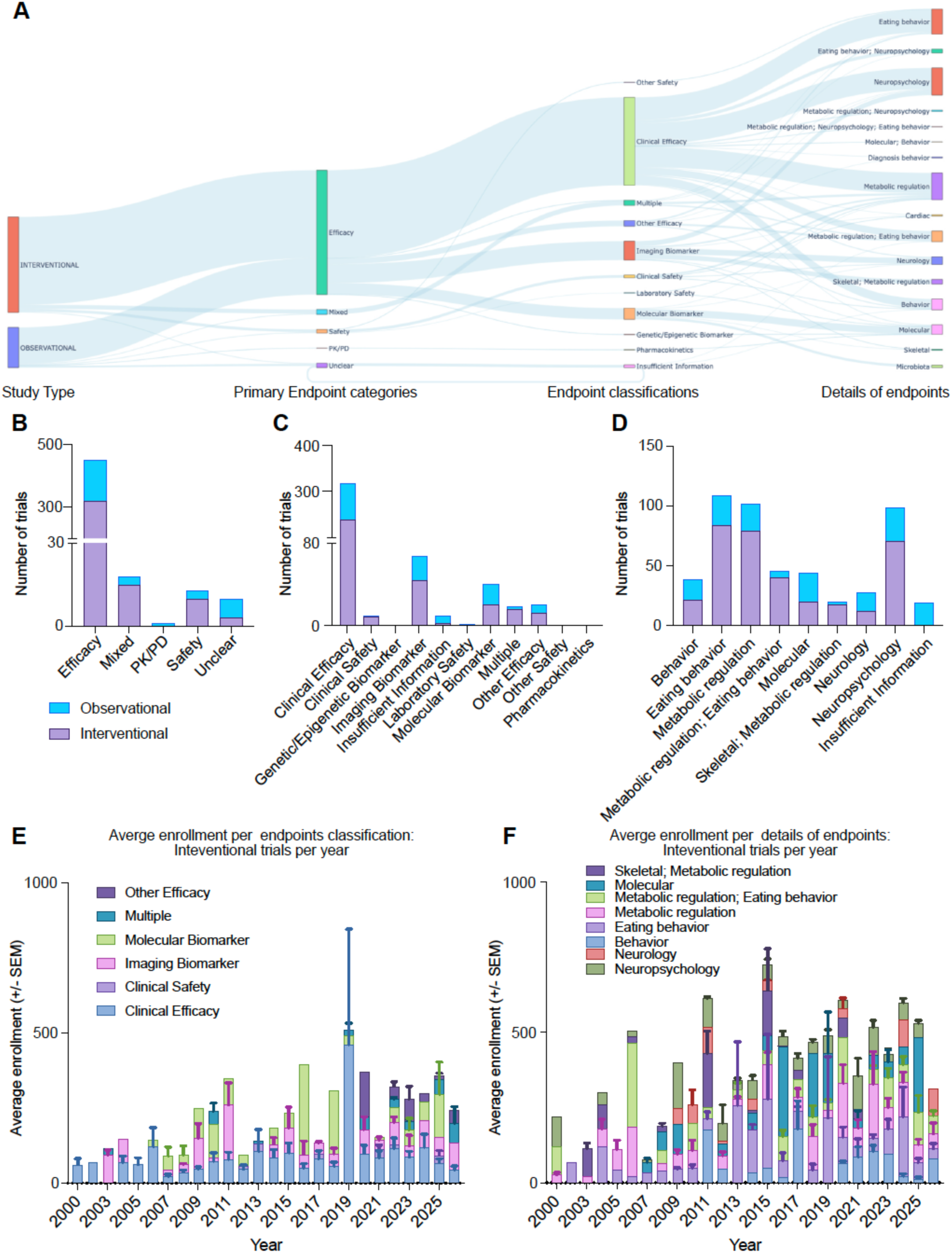
Comprehensive mapping and enrollment analysis of clinical trial endpoints in AN. (A) A sankey diagram illustrating the clusters and the hierarchical flow from study type (interventional vs. observational) to primary endpoint categories, endpoint classifications, and finally to highly specific pathophysiological and behavioral endpoint details. (B) Stacked bar chart detailing the total number of trials per primary endpoint category (Efficacy, Mixed, PK/PD, Safety, Unclear), stratified by study design. (C) Distribution of trials across distinct endpoint classifications, highlighting the prominence of clinical efficacy. (D) Quantitative breakdown of trials by specific endpoint details, such as eating behavior, metabolic regulation, and neuropsychology. (E) Longitudinal analysis (2000–2025) of average patient enrollment in interventional trials, categorized by broad endpoint classification. Error bars represent +/- SEM. (F) Longitudinal analysis (2000–2025) of average patient enrollment in interventional trials, categorized by specific endpoint details.

### Behavioral interventions and classic chemical drugs lead AN treatment strategy

An analysis of the intervention types utilized in AN clinical trial reveals a landscape heavily reliant on behavioral and pharmacological approaches (**Figure 5A, Supplementary table 1**). Behavioral interventions represent the largest single category, comprising 35.66% of trials, followed by drug therapies at 14.22%, and device-based interventions at 11.66% (**Figure 5A**). A large number of these interventions could not be categorized (denoted as “Others”, comprising 28.44%). When examining the underlying technological platforms for these drug therapies, “classic chemical drugs” constitute the vast majority of the research volume, significantly outpacing newer modalities like peptide therapeutics, monoclonal antibodies (mAbs) or microbiota-based treatments (**Figure 5B**).

**Figure 5.**
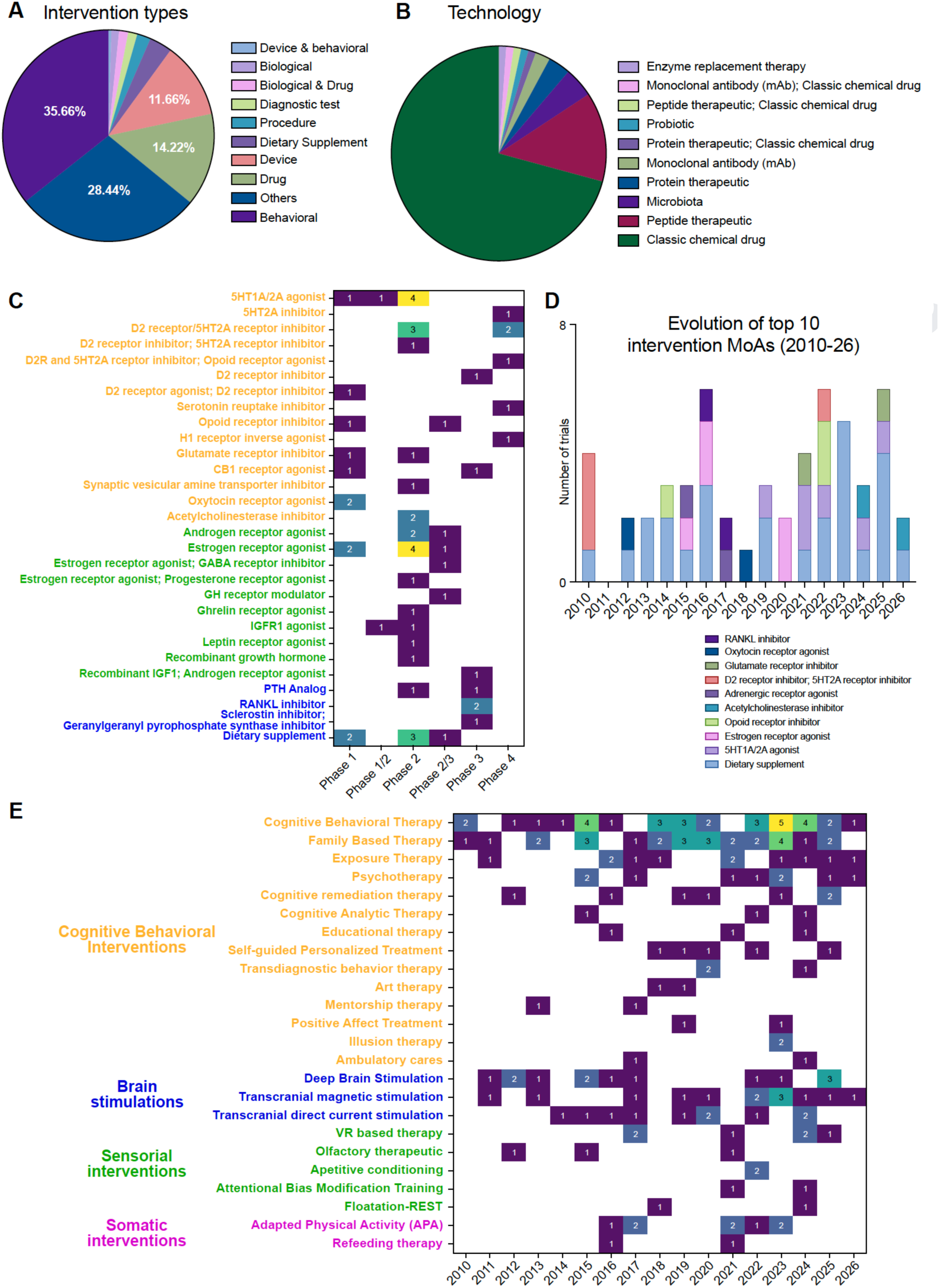
Intervention strategies and technological modalities in AN clinical trial. (A) Proportional distribution of broad intervention types, highlighting the dominance of behavioral, drug, and device-based approaches. (B) Breakdown of specific technological platforms utilized in drug-based interventions, illustrating the prevalence of classic chemical drugs. (C) Heatmap detailing the distribution of specific pharmacological mechanisms of action (MoA) across clinical trial phases (Phase 1 to Phase 4). (D) Longitudinal trends (2010–2026) in the number of trials employing specific receptor-targeted pharmacological interventions. (E) Heatmap illustrates the temporal utilization (2010–2026) of various non-pharmacological, behavioral, and device-based interventions.

A granular look at pharmacological mechanisms of action (MoA) across trial phases shows a highly fragmented landscape (**Figure 5C**). Most specific receptor targets (e.g., 5HT1A/2A agonists, Estrogen receptor agonists, D2 receptor inhibitors) are concentrated in Phase 2 trials, with very few advancing to Phase 3 or Phase 4. Longitudinally, trials utilizing dietary supplements and 5HT1A/2A agonists have seen consistent, albeit low-volume, investigation over the last decade. Significant neuromodulator therapies (such as 5HT1A/2A agonist, D2 receptor modulator, D2R and 5HT2A receptor inhibitor, CB1 receptor agonist, oxytocin receptor agonist, synaptic vesicle amine transporter inhibitor, etc.) have been evaluated as targeted therapies in AN (**Figure 5C, in dark yellow**), along with hormonal modulators for estrogen receptor, ghrelin, growth hormone (GH), androgen receptor, insulin like growth factor 1 (IGFR1) and others (**Figure 5C, in green**). PTH analog, RANKL inhibitor, sclerosin inhibitor and others have been evaluated in Phase 2 and Phase 3 trials for AN as well (**Figure 5C, in blue**). The longitudinal data from 2010 to 2026 highlights a dynamic shift in pharmacological priorities, where early-decade staples are being superseded by specialized bone-density and hormonal regulators (**Figure 5D**). Dietary supplements dominated the early landscape (2010–2015) but have seen a relative decline in trial frequency since 2020. Estrogen receptor agonists maintain a unique profile, remaining a top-10 MoA with a consistent trial presence that matches their high volume of Phase 2 activity (**Figure 5D**). Interestingly, while 5HT1A/2A agonists show the highest diversity across all phases (Phases 1, 2, and 4), their “top 10” peak was most pronounced between 2016 and 2019, suggesting a possible transition from high-volume broad testing to more targeted late-phase applications (**Figure 5D**). 5HT1A/2A agonist and acetylcholinesterase inhibitor show increased application since 2020 (**Figure 5D**).

The clinical trial landscape for non-pharmacological interventions between 2010 and 2026 is dominated by Cognitive Behavioral Interventions, with Cognitive Behavioral Therapy (CBT) maintaining the most consistent and high-volume presence, particularly peaking with 5 trials in 2023. Other psychological modalities such as Family Based Therapy and Exposure Therapy show steady longitudinal engagement, while newer formats like Self-guided Personalized Treatment established a consistent annual foothold starting in 2018 (**Figure 5E, in dark yellow**). In the realm of Brain Stimulations, activity intensified after 2011, with Transcranial Magnetic Stimulation (TMS) emerging as a leading modality reaching 3 trials in 2022, followed closely by steady research into Deep Brain Stimulation (DBS) and Transcranial Direct Current Stimulation (tDCS) (**Figure 5E, in blue**). Sensorial interventions reflect a more modern shift, specifically with the rise of VR based therapy trials appearing more frequently from 2022 onwards, alongside niche studies in Apetitive conditioning and Attentional Bias Modification Training (**Figure 5E, in green**). Finally, Somatic interventions are primarily represented by Adapted Physical Activity (APA), which saw distinct clusters of activity between 2016–2018 and 2021–2023, whereas Refeeding therapy remained a marginal area of focus with only sporadic trial entries (**Figure 5E, in pink**).

### Core pathophysiological endpoints drive research volume, with observational studies scaling massively

An analysis of the primary endpoints across the included clinical trials reveals that disease-specific psychopathology is the most frequently assessed outcome, followed closely by other clinical and psychiatric manifestations, and nutritional rehabilitation (**Figure 6A**). Further prominent endpoints encompass organ function and multi-system complications, as well as genetic, epigenetic, and neurobiological biomarkers. When examining the distribution of these trials by overarching research themes, the largest proportion is dedicated to combined metabolic regulation, which account for 20.08% of the total. This is followed closely by studies focusing on neuropsychological at 19.46% and eating behavior at 18.84%, and those investigating neurological aspects and brain metabolism at 8.07% (**Figure 6B**). The distribution also highlights emergence of composite primary endpoints such as metabolic regulation, neuropsychological, and eating behavior studies and neurological implications accounting for almost 12% of the trial endpoints. Assessment of average participant enrollment highlights stark contrasts between interventional and observational study designs across various research domains. Observational studies exhibit the largest cohort sizes, most notably within trials focusing on combined metabolic regulation and eating behavior, which average 1992.33 participants (**Figure 6C**). Similarly, large observational cohorts are seen in molecular studies with an average enrollment of 1586.72, and isolated eating behavior trials averaging 1167.84 participants (**Figure 6C**). Conversely, interventional studies generally feature much smaller sample sizes, with the highest average enrollment observed in combined eating behavior and neuropsychological trials at 574.08 participants. Other domains, such as cardiac research and combined skeletal and metabolic regulation studies, maintain relatively restricted cohort sizes across both design types, typically remaining well below 220 participants (**Figure 6C**).

**Figure 6.**
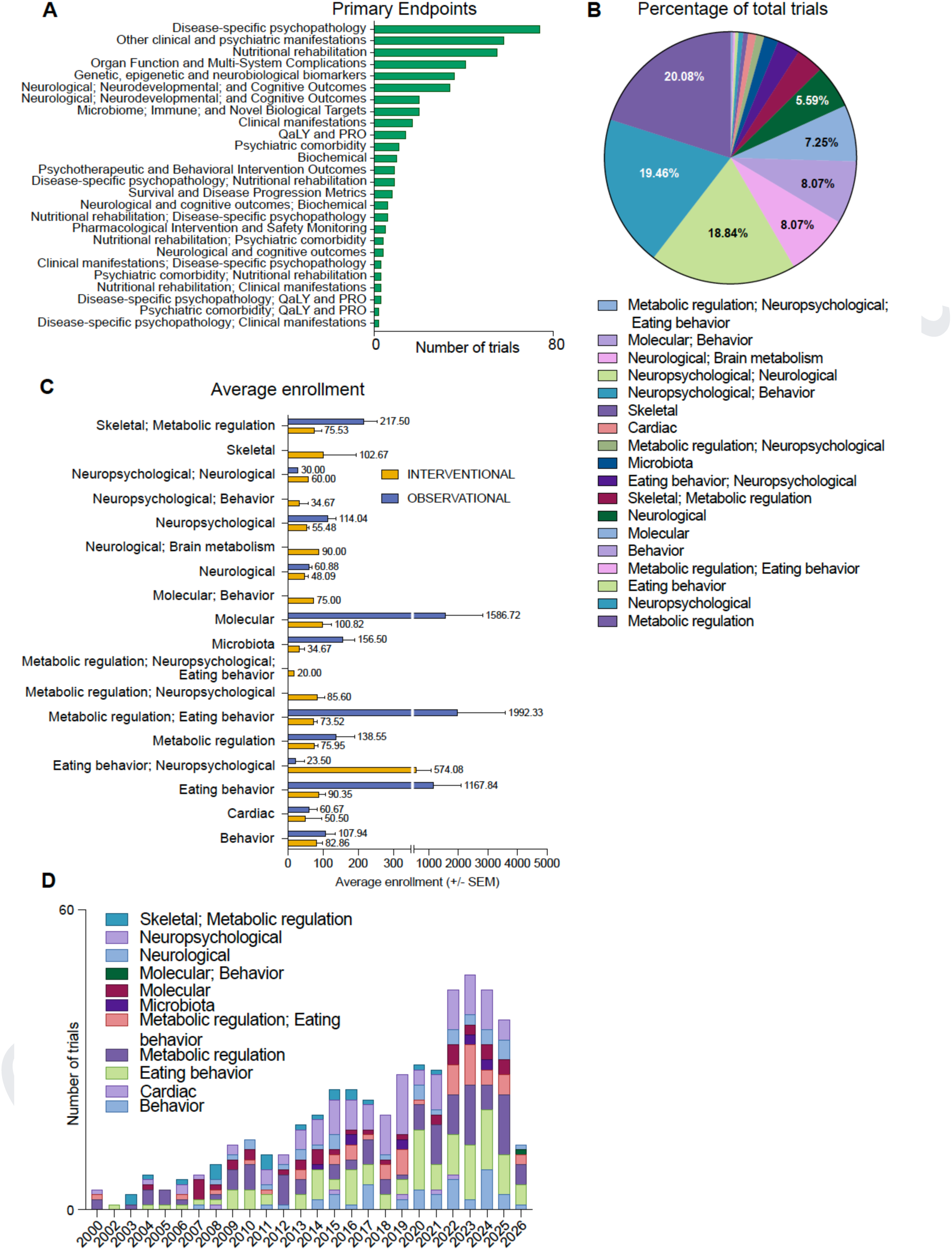
Comprehensive analysis of primary endpoints and associated patient enrollment in AN clinical trial. (A) Distribution of clinical trials across diverse primary endpoint categories, emphasizing disease-specific psychopathology and clinical manifestations. (B) Proportional distribution of the total trial landscape categorized by key pathophysiological and behavioral domains. (C) Comparison of average patient enrollment between interventional and observational study designs, stratified by specific endpoint clusters. (D) Longitudinal analysis spanning 2000 to 2026, tracking the annual volume of trials categorized by these key domains.

Finally, longitudinal tracking of trial initiation from the year 2000 onward demonstrates a substantial and sustained expansion in the overall volume of clinical research within this field (**Figure 6D**). The early 2000s were characterized by a relatively sparse number of trials, but a clear upward trajectory in research activity began around 2013. This growth accelerated significantly over the subsequent decade, culminating in peak trial volumes between 2022 and 2024 (**Figure 6D**). This temporal increase is characterized by a diversification in the types of trials conducted, with sustained and growing contributions from studies focused on eating behavior, metabolic regulation, and complex neuropsychological interactions driving the recent surge in annual trial counts.

### Anorexia trial complexities are highlighted by clustering of adjacent indications, highlighting complex and overlapping endpoints

The analysis of the co-occurrence network reveals that AN trials clusters with other major eating disorders and physiological comorbidities and adjacent indications (**Figure 7A**). The strongest association is with the general term “eating disorders,” which accounts for 102 co-occurrences, followed by a significant overlap with “bulimia nervosa” at 67 instances and “binge-eating disorder” at 31 instances (**Figure 7B**). This indicates a high level of diagnostic fluidity or concurrent reporting within the spectrum of eating pathology. Beyond these primary links, the data highlights a clear developmental focus, with “Atypical AN” appearing 19 times and “Anorexia in Adolescence” and “AN Restricting Type” each appearing 10 times. The results highlight the systemic physical toll of the disorder, mapping direct connections to bone health issues such as “Osteoporosis” (7) and “Bone Loss” (3), alongside psychological comorbidities including “Anxiety” (7), “Depression” (6), and “Panic Disorder”. Furthermore, the network captures specific clinical states, such as “AN in Remission” (4) and “Avoidant Restrictive Food Intake Disorder” (3), illustrating the complex, multi-faceted nature of the condition’s clinical presentation (**Figure 7B**).

**Figure 7.**
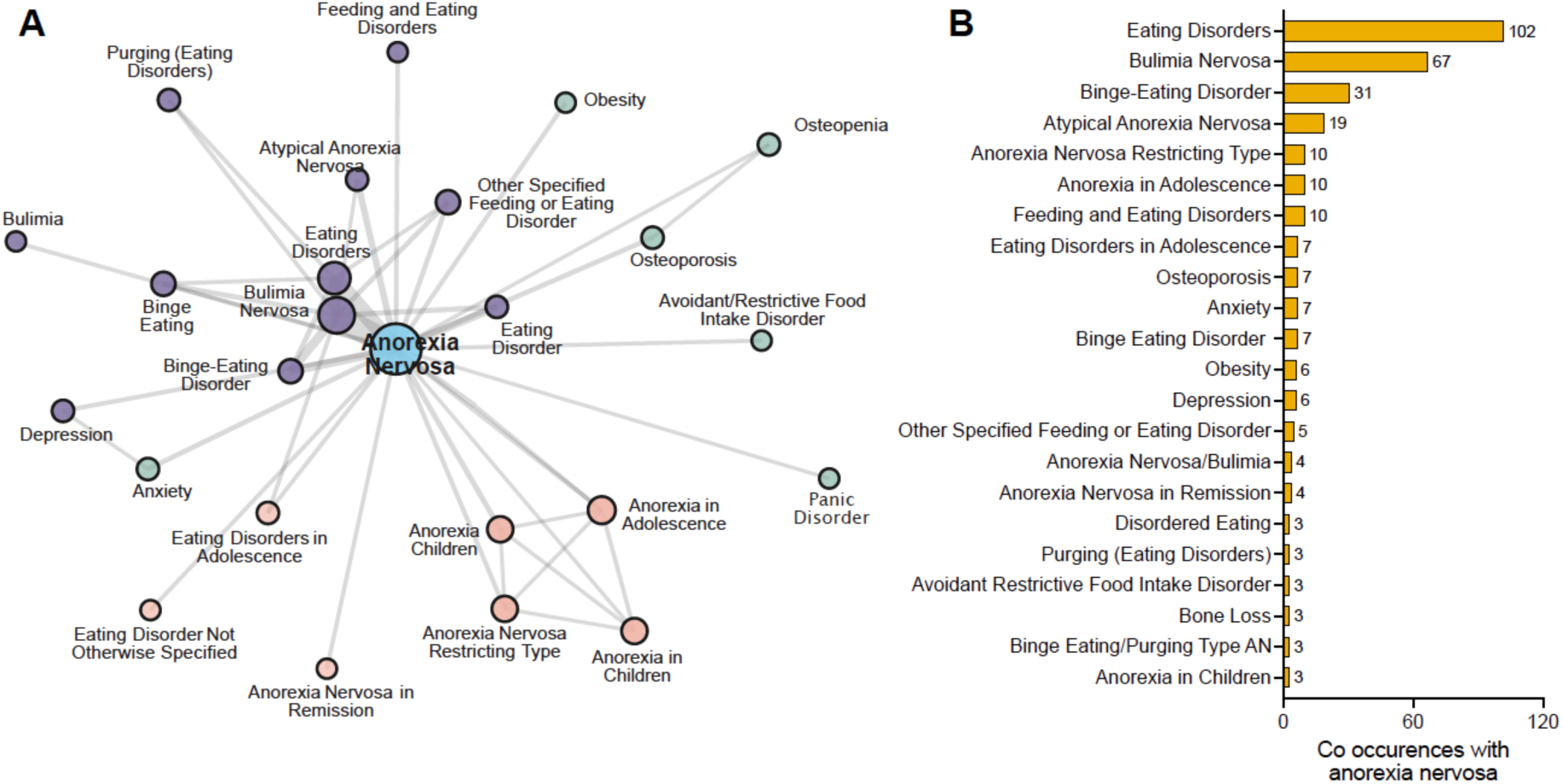
Co-occurrence analysis of AN with related clinical conditions and terms. (A) A network map illustrating the connectivity between AN (central node) and various feeding, eating, and comorbid disorders. The distance and line thickness represent the strength of association between terms. (B) A bar chart quantifying the specific number of co-occurrences between AN and the top associated clinical terms. Values at the end of each bar indicate the absolute count of documented co-occurrences.

## Discussion

Over the past 25 years, nearly 400 studies have investigated AN within the framework of neurobiological and neurocognitive research, highlighting the sustained scientific interest in understanding the mechanisms underlying this disorder. The increasing number of studies registered over time reflects ongoing efforts to better characterize the pathophysiology of AN and to develop new therapeutic strategies.

Approximately 71% of the identified studies were classified as interventional. However, this finding should be interpreted with caution. According to the National Institutes of Health, a clinical trial is defined as any research study in which human participants are prospectively assigned to one or more interventions to evaluate health-related outcomes. This definition includes not only therapeutic interventions but also diagnostic procedures, imaging protocols, and mechanistic experimental manipulations (National Institutes of Health. Definition of a Clinical Trial; NIH Clinical Trials Definition and Classification). Consequently, studies involving the administration of biological tracers, neuroimaging probes, or physiological manipulations may be classified as interventional trials even when their primary objective is exploratory rather than therapeutic. Regulatory initiatives such as the Final Rule governing ClinicalTrials.gov reporting have further expanded the scope and transparency requirements for registered trials (Zarin et al., 2016).

Among trials for which phase information was available, phase II studies represented the largest category. However, approximately 75% of studies were not classified according to conventional trial phases (phase I–III). This apparent discrepancy further supports the interpretation that many of the studies registered as interventional correspond to exploratory investigations aimed at better understanding disease mechanisms rather than classical drug development programs.

Geographically, the distribution of studies revealed a strong predominance of the United States, followed by France and Italy. In contrast, Asian countries and Russia were markedly underrepresented. This finding may reflect the use of region-specific clinical trial registries in Asia that are not systematically captured in ClinicalTrials.gov. Similarly, studies conducted in Russia were not represented in the database analyzed. Overall, these results highlight the strong predominance of North American research activity in this field. However, this geographic distribution should be interpreted with caution, as our analysis relied on ClinicalTrials.gov, which may underrepresent studies conducted in Asia or Russia. Several countries use independent national registries - such as the Chinese Clinical Trial Registry, the Clinical Trials Registry of India, or the Japanese Registry of Clinical Trials - which are not always systematically indexed in ClinicalTrials.gov, potentially leading to regional registration bias.

Another notable finding was the relatively high proportion of terminated or discontinued studies, reaching approximately 45%. Although the COVID-19 pandemic may have contributed to some of these interruptions, the temporal distribution of trial discontinuations suggests that additional factors may be involved. One likely explanation is the difficulty in recruiting patients with AN into interventional studies.

Recruitment challenges in randomized clinical trials involving patients with AN have been widely documented in the literature. Several methodological reports have highlighted the difficulties associated with enrolling participants in multicenter trials, often due to strict inclusion criteria, limited patient availability, and the complexity of trial designs (Lock et al., 2012). Earlier studies examining recruitment in AN trials also reported substantial challenges related to patient engagement and retention (McDermott et al., 2004).

In addition to methodological constraints, patient-related factors may also contribute to recruitment difficulties. Individuals with AN often experience ambivalence toward treatment and recovery, which may influence their willingness to participate in experimental interventions. Qualitative research exploring patient perspectives on treatment has shown that individuals with AN frequently expect therapeutic approaches to address not only weight restoration but also psychological and social aspects of the disorder. These expectations, combined with concerns about loss of control or treatment coercion, may influence participation in clinical research.

The differences observed in sample sizes between observational and interventional studies further support this interpretation. Observational studies tended to include substantially larger populations than interventional trials. While this difference may partly reflect retrospective study designs, it may also indicate that observational protocols are easier to implement within routine clinical care settings compared with controlled interventional trials.

The limited number of trials targeting specific biological pathways is also noteworthy. Over the past decades, numerous studies have investigated circulating peptides and neuroendocrine signals involved in appetite regulation, metabolic adaptation, and energy homeostasis in AN. However, only a small proportion of these candidate mechanisms appear to have progressed toward structured clinical trial programs. This observation echoes findings from recent scoping analyses of peptide-based biomarkers and regulatory pathways in AN, which highlighted a substantial gap between mechanistic discoveries and clinical development.

Qualitative research has provided valuable insights into how individuals with AN perceive treatment and engage with care. Interview-based studies have shown that patients often expect treatment not only to promote weight restoration but also to address cognitive, emotional, and social difficulties associated with the disorder (Paulson-Karlsson and Nevonen, 2012). Other qualitative investigations have explored barriers and facilitators to treatment initiation, highlighting the importance of psychological ambivalence, perceived treatment efficacy, and the quality of the therapeutic relationship (Kästner et al., 2021). Studies examining the reasons for accepting or declining treatment further indicate that fears of losing control, identity-related concerns, and apprehension about coercive care may influence treatment decisions (Andersen et al., 2021; Gorse et al., 2013). Finally, qualitative work on recovery experiences emphasizes the importance of autonomy, trust in clinicians, and individualized care in sustaining treatment engagement. Together, these findings suggest that patient perceptions and expectations play a central role in engagement with treatment and may partly explain the recruitment and retention difficulties observed in clinical trials in AN.

When considering research objectives, the majority of studies focused on clinical efficacy, both in interventional and observational designs. Three main research domains emerged from the analysis: eating behavior, neuropsychological mechanisms, and metabolic regulation. Secondary domains included imaging biomarkers and clinical safety outcomes. Interestingly, despite the large proportion of studies classified as interventional, relatively few specifically addressed safety outcomes, again suggesting that many trials may primarily pursue exploratory or mechanistic objectives rather than classical therapeutic development pathways.

Temporal trends revealed a relatively stable number of studies investigating clinical efficacy over the 25-year period, with no evident decrease during the COVID-19 pandemic. In contrast, a noticeable increase in studies categorized as “multiple-domain” investigations have been observed since approximately 2019. These studies typically combine several biomarkers or multimodal assessments, suggesting a gradual shift toward more integrative and multidimensional research strategies.

Regarding the types of interventions investigated, behavioral interventions and pharmacological treatments each accounted for approximately one third of interventional studies. Behavioral interventions were dominated by cognitive-behavioral therapy and family-based therapy. The high representation of family-based and cognitive-behavioral interventions appears consistent with current clinical recommendations. Recent clinical guidelines, including the 2023 American Psychiatric Association practice guideline and the NICE guideline, recommend family-based treatment as the first-line intervention for adolescents with AN and specialized psychotherapeutic approaches for adults, including cognitive-behavioral therapy among the preferred treatment options (Crone et al., 2023; *National Institute for Health and Care Excellence*, 2020).

Conversely, relatively few studies investigated innovative biological approaches such as peptide-based therapies, immunomodulatory strategies, or microbiota-related interventions. This observation is somewhat surprising given previous evidence highlighting the potential importance of circulating peptides in AN. In a recent scoping review, our group identified 207 peptides reported in the literature in relation to AN (Doua et al., 2025). The limited translation of these findings into clinical trials likely reflects the considerable heterogeneity in peptide measurement techniques, phenotypic characterization, and study designs, which complicates the generation of robust therapeutic hypotheses.

Among pharmacological targets, serotonergic pathways and the gonadotropic axis were the most frequently investigated. Studies targeting serotonergic mechanisms remained relatively stable over time, reflecting the long-standing interest in serotonin dysfunction in AN. In contrast, research focusing on the gonadotropic axis reflects the clinical importance of endocrine complications associated with chronic undernutrition, including hypothalamic hypogonadism and bone health alterations frequently observed in patients with AN.

Several limitations should be considered when interpreting the present findings. First, the analysis relied exclusively on ClinicalTrials.gov, which may not capture all clinical studies conducted worldwide. Countries that rely primarily on national trial registries may therefore be underrepresented. Second, registry entries may contain incomplete or inconsistently updated information regarding study design, recruitment status, or outcomes. Despite these limitations, clinical trial registries provide an important complementary perspective on research activity by capturing ongoing studies, discontinued trials, and emerging research directions that may not yet appear in the published literature.

Taken together, these findings highlight the considerable scientific effort devoted to AN research over the past decades while simultaneously revealing important gaps between mechanistic knowledge and therapeutic development. Understanding how biological insights are translated into clinical research programs may represent a key step toward accelerating the development of effective treatments for this severe and still largely unmet medical condition.

## Data Availability

The analytical pipeline in this study was executed using ClinForecast(TM). Because this tool is a protected commercial product, the associated source code and proprietary algorithms are strictly confidential and are not available for public sharing. However, the aggregated datasets generated during the current study are available in the results and supplementary materials. The dataset can be found in Supplementary table 1.

## Declarations

### Ethics declaration

Not applicable as no human samples or human derived datasets were processed.

### Competing interests

The authors declare that they have no competing interests. Manish Sarkar and Dimitrije Milunov are employees of MedInsights SAS. Soham Saha is the founder and CEO of MedInsights SAS. Dr. Kingsley Urakpo is a medical advisor and board member to MedInsights SAS and owns NADDBio LLC.

### Code and data sharing

The analytical pipeline in this study was executed using ClinForecast™. Because this tool is a protected commercial product, the associated source code and proprietary algorithms are strictly confidential and are not available for public sharing. However, the aggregated datasets generated during the current study are available in the results and supplementary materials. The dataset can be found in Supplementary table 1.

### Author contributions

Bo.Ga, N.G. and S.S. led the study, including ideation, conceptualization, study design, analytical pipeline, data analysis, interpretation and writing the manuscript. M.K. and D.M. contributed to the study design, computational analysis and supported data interpretation. M.S. contributed to developing the theoretical framework, conducting quality checks and ensured completeness. Bé.Ga conducted initial model training on ClinForecast^TM^. K.U. read the manuscript draft and suggested changes. All authors revised, reviewed and approved the final manuscript.

## Abbreviations

5HT1A/2A: Serotonin 1A/2A receptor
AAV: Adeno-associated virus
AN: Anorexia nervosa
APA: Adapted physical activity
API: Application Programming Interface
BEST: Biomarkers, Endpoints, and other Tools
BMI: Body mass index
CAR-T: Chimeric antigen receptor T cells
CBT: Cognitive Behavioral Therapy
CRP: C-reactive protein
CTX: C-telopeptide of type I collagen
DSM-5: Diagnostic and Statistical Manual of Mental Disorders, Fifth Edition
GPT: Generative pre-trained transformer
ICTRP: International Clinical Trials Registry Platform
IL-6: Interleukine 6
LLM: Large language model
mAbs: Monoclonal antibody
MoA: Mechanism of Action
MRI: Magnetic resonance imaging
P1NP: Intact N-Terminal Propeptide of Type I Procollagen
PRO: Patient reported outcomes
PROTAC: Proteolysis-targeting chimera
PTSD: Post traumatic stress disorder
QaLY: Quality adjusted life years
TNF-α: Tumor necrosis factor alpha
xAI: Explainable AI

## Notes

### Funding Statement

This study did not receive any funding

